# A longitudinal seroprevalence study in a large cohort of working adults reveals that neutralising SARS-CoV-2 RBD-specific antibodies persist for at least six months independent of the severity of symptoms

**DOI:** 10.1101/2020.12.22.20248604

**Authors:** Angelika Wagner, Angela Guzek, Johanna Ruff, Joanna Jasinska, Ute Scheikl, Ines Zwazl, Michael Kundi, Hannes Stockinger, Maria R. Farcet, Thomas R. Kreil, Eva Hoeltl, Ursula Wiedermann

**Author notes:** Correspondence to: Prof. Ursula Wiedermann, MD, PhD, Institute of Specific Prophylaxis and Tropical Medicine, Centre for Pathophysiology, Infectiology and Immunology, Medical University of Vienna, Kinderspitalgasse 15, Vienna A-1090, Austria.

## Abstract

**Background:** In spring 2020, at the beginning of the first pandemic severe acute respiratory syndrome coronavirus 2 (SARS-CoV-2) wave in Europe, we set up an assay system for large-scale testing of virus-specific and protective antibodies including their longevity.

**Methods:** We analysed the sera of 1655 adult employees for SARS-CoV-2-specific antibodies using the S1 subunit of the spike protein of SARS-CoV-2. Sera containing S1-reactive antibodies were further evaluated for receptor-binding domain (RBD)- and nucleocapsid protein (NCP)-specific antibodies in relation to the neutralisation test (NT) results at 0, three and six months.

**Findings:** We found immunoglobulin G (IgG) and/or IgA antibodies reactive to the S1 protein in 10.15% (n=168) of the participants. In total, 0.97% (n=16) were positive for S1-IgG, 0.91% (n=15) were S1-IgG-borderline and 8.28% (n=137) exhibited only S1-IgA antibodies. Next, we evaluated the 168 S1-reactive sera for RBD- and NCP specificity: 8.33% (n=14) had detectable RBD-specific and 6.55% (n=11) NCP-specific antibodies. The latter correlated with NTs (kappa coefficient = 0.8660) but started to decline already after 3 months. RBD-specific antibodies correlated best with the NT (kappa = 0.9448) and only these antibodies were stable for up to six months. All participants with virus-neutralising antibodies reported symptoms, of which, anosmia and/or dysgeusia correlated best with the detection of virus-neutralising antibodies.

**Interpretation:** RBD-specific antibodies were most reliably detected post infection, independent of the number/severity of symptoms, and correlated best with protective neutralising antibodies at least for six months. They thus qualify best for large-scale seroepidemiological evaluation of both seroprevalence and seroprotection.

**Funding:** This study received funding from the Austrian Ministry of Education, Science and Research within the research framework in relation to the coronavirus disease 2019 pandemic (GZ 2020 0225 104).

**Key points:** Persistence of SARS-CoV-2 antibodies depends on their specificity. Total RBD-specific antibodies are those that are stable for up to at least six months and correlate best with neutralisation independent of the presence and severity of COVID-19 symptoms.

**Research in context:** *Evidence before the study:* At the beginning of the study (early pandemic in April 2020), the SARS-Cov-2 specific seroprevalence was totally unknown. Additionally, S1-specific antibody assays being the first on the market were tested with limited sample size showing a lower sensitivity and specificity at that time. Furthermore, at that time, there were no unambiguous interpretations of antibody test results with regard to immunity/protection against reinfection. It was also not clear whether the detection of different antibody specificities could yield an essential input into the interpretation of the antibody’s qualities. Another open question was how long antibodies of the various specificities as well as antibodies with protective capacities would persist.

*Added value of this study:* We provide data to confirm the most reliable correlation of RBD-specific antibodies with neutralising antibodies that are stable for at least six months. S1- and NCP-specific antibodies wane more quickly than RBD-specific antibodies, rendering them not as ideal candidates for longitudinal seroprevalence studies. Concerning symptoms, anosmia/dysgeusia was strongly associated with NT-seropositivity and seroprotection in the overall study population.

*Implications of all the available evidence:* Our data suggest that RBD-specific total antibody measurements with assays of high specificity can be used for cross-sectional as well as longitudinal seroepidemiological studies, even in low-prevalence settings. Detection of these antibodies also indicates robust seroprotection for at least six months. Due to the substantial loss of S1- and NCP-specific antibodies within the first months, assays targeting these antigen specificities – in contrast to RBD-specific antibody measurements – are not optimal to assess the duration of seroprotection. Overall, respiratory symptoms alone were not useful in predicting a past infection with SARS-CoV-2. However, anosmia/dysgeusia appeared to be a significant diagnostic marker, in particular for mild COVID-19.

## Introduction

The ongoing severe acute respiratory syndrome coronavirus 2 (SARS-CoV-2) pandemic has led to dramatic restrictions in public life worldwide to mitigate the anticipated epidemic peak.^1^ At the early stage of the pandemic, data were missing to estimate the actual number of infected individuals, including asymptomatic/oligosymptomatic cases that were left unreported due to the limitations in the polymerase chain reaction (PCR) testing capacity and strategy that was initially confined to the fulfilment of case definitions. It was, therefore, difficult to assess the actual risk of infection for employers with respect to shared workspaces and staff in contact with customers. Where possible, employers facilitated employees’ work in home office mode with the intention to reduce the number of social contacts and consequently, the risk of infection.^2^

In order to estimate past infections irrespective of symptoms and a preceding PCR test, specific and sensitive serological antibody tests, including neutralising assays, are necessary tools.^3^ Additionally, serological tests are an important instrument for epidemiological surveillance and to assess whether/when herd immunity has been reached within a population.

Several validated serological formats are now in use, based on enzyme-linked immunosorbent assays (ELISA) and chemiluminescence targeting different SARS-CoV-2 antigens.^4^ The target antigens are the spike (S) protein with its receptor-binding domain (RBD), and the nucleocapsid protein (NCP). Both antigens have been shown to induce robust antibody responses in infected individuals.^5,6^ Another important question has emerged as to whether the detected antibodies also indicate protection against reinfection. In this regard, testing for neutralising antibodies is considered to be the gold standard as it additionally assesses antibody functionality, i.e. virus inactivation.^6^ However, such testing is laborious, time-consuming and requires biosafety level three laboratory space, and therefore not suitable for large-scale testing/seroprevalence surveys. Along these lines, it is still unclear which test would prove most appropriate to describe transmission patterns and to determine immunity upon virus contact in overall as well as in defined populations (in contrast to individual analysis/neutralisation tests [NTs]).

With the beginning of an exponential increase in coronavirus disease 2019 (COVID-19) cases in Austria, the government declared a public lockdown with March 16^th^, 2020. At this time, the actual rate of infections was unknown and consequently, the risk of virus exposure for personnel in various work settings.

The goal of the current study was to analyse the seroprevalence of SARS-CoV-2-specific antibodies at the beginning of the pandemic and over several months in a representative cohort of employees from a large Austrian company. Besides, the evaluation of seroprotection and persistence of protective antibodies were /critical aspects of this study for which several assays were included to identify the most accurate test for large-scale seroepidemiological analysis. The participants were included irrespective of a previous history of COVID-19 or experienced symptoms. Already before study onset, the employees were split into two groups, those sent into home office and those remaining on-site. The accumulated basic demographic data (gender, age, household size) and information on respiratory tract infections and symptoms, medical risk factors and travel history were analysed in context with the SARS-CoV-2-specific antibody test results. Employees with virus-reactive antibodies at the initial blood draw were invited for follow-up blood draws at three and six months after study onset to analyse the persistence of the detected antibody levels. At six months, also participants without detectable virus-reactive antibodies at the initial blood draw were asked for a follow-up blood draw in order to detect seroconversion and to assess the development of seroprevalence in the overall study population over the last months.

## Methods

### Patients and samples

We included 1655 serum samples of employees working for a large company in Vienna. While half of the staff continuously worked on-site with frequent client contacts, the other half worked from home at the beginning of this trial followed by a weekly rotation between home office and on-site work after the lockdown period in Austria. The blood samples were taken at the medical centre of the company between 2^nd^ and 17^th^ April 2020 and sent in for further analysis (to the Institute of Specific Prophylaxis and Tropical Medicine at the Medical University of Vienna. The employees gave informed consent to SARS-CoV-2 serological testing and answered a questionnaire covering demographic data and their medical history, including current medications (see Supplement materials). The ethics committee of the Medical University of Vienna approved this monocentric study (EK 1438/2020, EK 1746/2020).

### Testing for SARS-CoV-2 antibodies

The SARS-CoV-2 specific antibody levels were measured with four different serological assays. After determining S1-specific IgG and IgA by ELISA, we further analysed those presenting positive or borderline results for RBD- and NCP-specific antibodies and by a live virus neutralisation test (at Takeda’s Global Pathogen Safety unit in Vienna) (for detailed information see supplementary materials).

### Statistical evaluation

Data were evaluated for the two groups (working on-site and from home) and the subgroups stratified for age (15 to 25 years, 25 to 50 and above the age of 50). Seropositivity as a dependent variable was evaluated in a general linear model for binominal counts (see supplementary materials for more details).

## Results

### Study populations, demographic data

Overall, the study population consisted of 1655 volunteers, with an almost balanced female/male ratio (53.53%/46.47%) (table 1, suppl. figure 1). The majority of the study population (62.30%) was in the medium-age group (25-50 years). Higher percentages of the medium-age and older age groups (53.83% and 62.78%, respectively) had been sent home to work in contrast to only 27.88% of the young age group. In relative terms, 52.33% of the total study population worked from home. Less than half of the participants (42.05%) reported that they had experienced symptoms compatible with COVID-19 within the last three months before the beginning of the study, with cough, rhinitis, sore throat, and fever being most frequently mentioned (suppl. figure 2a). One quarter (23.63%) had risk factors in their medical history, which potentially predisposed for a severe COVID-19 disease (suppl. figure 2b). However, 145 (8.76%) did not give details about the type of risk factors. Furthermore, 24.35% reported regular medication uptake (table 1, suppl. figure 3).

**Table 1:**
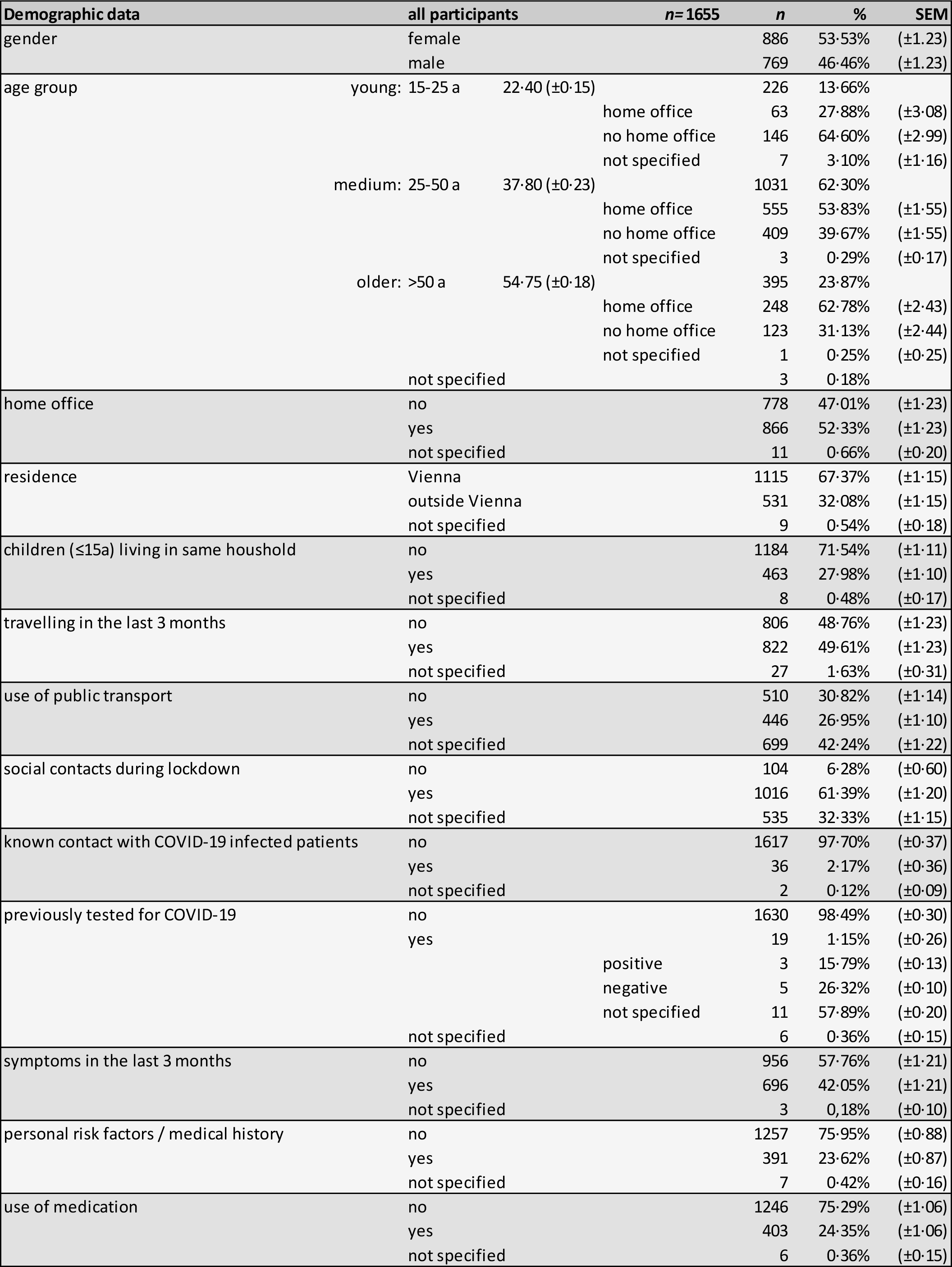
Demographic data by general characteristics brief medical history. These data were analysed from the received questionnaires. The different demographic parameters are

### Sample analyses

Serological testing for SARS-CoV-2 S1-specific IgG and IgA antibodies revealed that 1487 participants (89.85%) were seronegative (figure 1a). Thus, 10.15% displayed S1-reactive antibodies, among whom 16 (9.52%) were IgG-positive, 15 (893%) IgG-borderline, and 137 (81.55%) exhibited only IgA antibodies without IgG antibodies.

**Figure 1.**
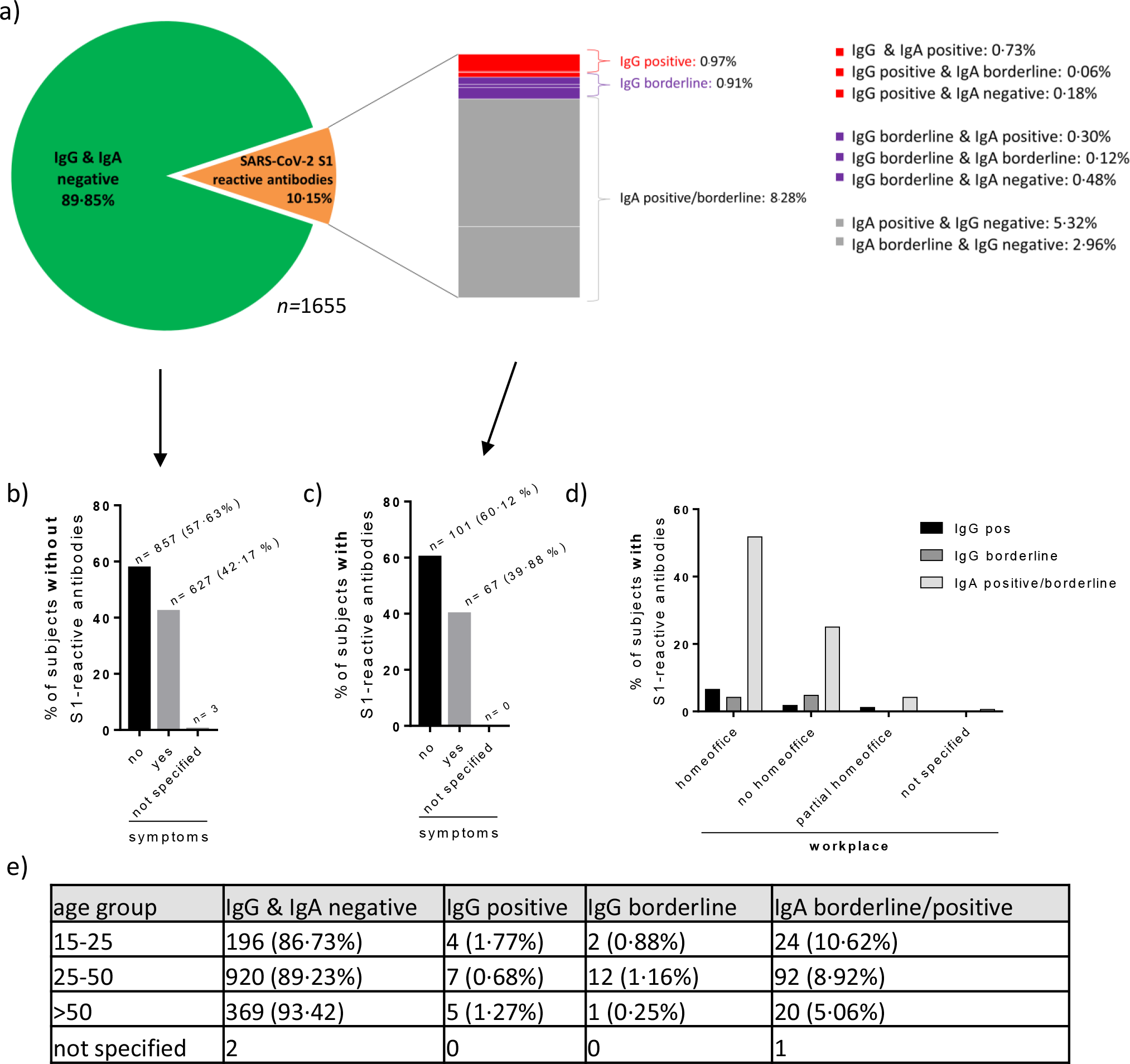
S1-reactive IgA and IgG antibody results presented as proportions of the total study population (a), of those without (b) and with detectable S1-reactive antibodies (c) dependent whether symptoms were recorded, according to workplace (d), and age group (e). Antibodies were measured in sera from the initial blood draw at day 0.

Further description of participants with antibodies reactive to S1 showed that COVID-19-associated symptoms were recorded in 39.88% of the participants with and in 42.17% without S1-reactive antibodies (figures 1b and c). With respect to the working situation, the highest rate of S1-positive or -borderline participants was home office workers before and during the beginning of the study (figure 1d). Regarding age, the highest number of participants with virus-reactive antibodies were between 25 and 50 years old (medium-age group) (n=111, 6.71% of all participants) (figure 1e).

The statistical analysis of the demographic and medical data revealed that three factors significantly correlated with S1 seropositivity (table 2), i.e. (i) age, decreasing seropositivity with increasing age, (ii) home office, and (iii) loss of taste and/or smell. Concerning symptoms, anosmia/dysgeusia showed the highest probability for a positive virus (S1)-specific antibody result (odds ratio 22.48), whereas the presence of any other reported symptom did not.

**Table 2:** Probability of antibody positivity according to independent variables (predictors) such as home office, known contact with COVID-19 patients, children < 15 years of age within the same household, residence in Vienna or outside, age group (comparison to the young 15- to 25-years-old group) and the symptoms of anosmia and/or dysgeusia.

| **predictor** | **Odds ratio** | **95% CI** | **p - value** |
| --- | --- | --- | --- |
| **home office (y/n)** | **1·91** | **1·21-3·01** | **0·005** |
| **Known contact with COVID-19 patients (y/n)** | 1·27 | 0·35-4·59 | 0·719 |
| **Children < 15y in the same household (y/n)** | 1·04 | 0·65-1·64 | 0·882 |
| **Residence in Vienna (y/n)** | 1·32 | 0·86-2·03 | 0·197 |
| **Age group 15-24** | 1·00 | - | - |
| **Age group 25 -50** | **0·56** | **0·32-0·99** | **0·046** |
| **Age group >50** | **0·38** | **0·19-0·77** | **0·007** |
| **Anosmia/dysgeusia** | **22·48** | **7·75-65·22** | **<0·001** |

To further characterise antibody responses, all 168 sera with detectable S1-specific antibodies were also tested for RBD- and NCP-specific antibodies. Only a part of the S1-reactive sera also displayed RBD- or NCP-specific antibodies: 8.33% (14/168) had detectable RBD-specific total antibodies, whereas ten of these sera were also positive for S1-specific IgG, with two IgG borderlines and two IgG negatives (figure 2). The NCP-specific antibodies were positive in 6.55% (11/168) of the respondents with S1-reactive antibodies, with seven also being IgG-positive, one IgG-borderline and two IgG negatives. Of the S1-IgA-positive or -borderline samples, two were positive for RBD and two for NCP (one of these for both).

**Figure 2.**
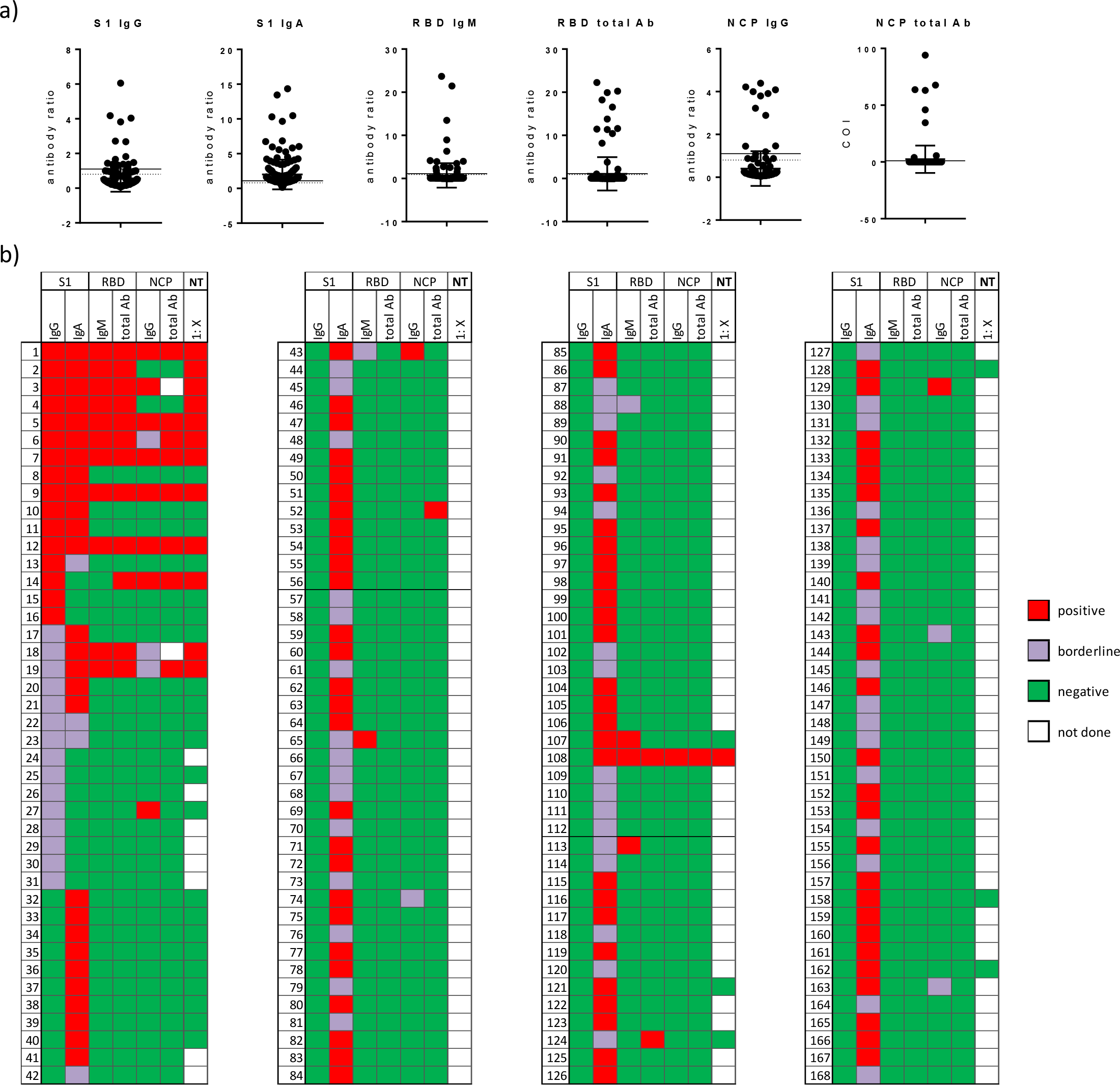
Individual antibody results against different SARS-CoV-2 antigens expressed as antibody ratio or cutoff index (COI) (a) and as cumulative results (b) of all participants with S1-reactive antibodies (n=168) at the initial blood draw (day 0). Antibody results: red = reactive; violet = borderline; green = not reactive; white = n.d. (=not done). Antibody (Ab)

Virus-neutralising antibodies are regarded as surrogate of protection. For detection of these functional antibodies within the group of S1-positive sera, the NT, regarded as the gold standard among the SARS-CoV-2-specific serological assays, was performed. The results revealed that ten out of 16 S1-IgG-positive, two out of nine S1-IgG-borderline and only one of the S1-IgA positive/borderline sera were also positive in the NT (figure 2, suppl. figure 5).

We then analysed whether the antibodies directed against the various SARS-CoV-2 antigens, such as S1, RBD or NCP, correlated with the presence of neutralising/protective antibodies. The RBD-specific antibodies showed the highest agreement with the NT – total antibodies, kappa (k)=0.94; p<0.0001 and IgM (k=0.89; p<0.0001) (figure 3), indicating that these RBD-specific antibodies can be used as a predictor/surrogate for antibodies with protective properties. By contrast, S1-specific IgA failed to show any correlation with the neutralising antibody levels. Furthermore, the quantity (antibody level) of S1-specific IgG and IgA was not predictive for a positive NT result (suppl. figure 5a and b).

**Figure 3.**
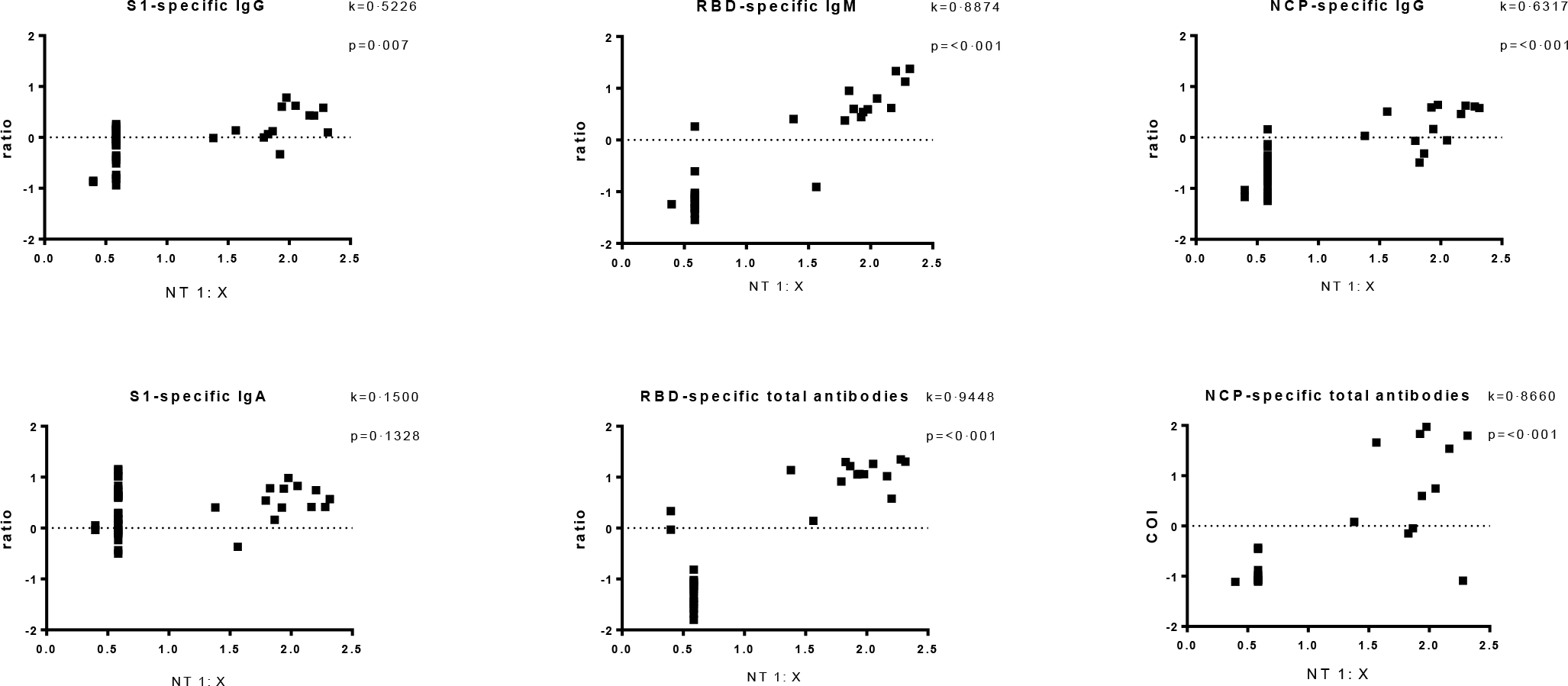
Correlation of SARS-CoV-2-specific antibody test results expressed as antibody ratios or cutoff index (COI) with the neutralisation test (NT) result expressed as titer 1:X measured in sera from the initial blood dray at day 0. Antibody ratios, COI and NT titers after logarithmic transformation. Kappa coefficient indicating agreement between the assays (very good > 0·8). The results presented as ratio or COI depend on the manufacturer’s instructions.

All participants with neutralising antibodies reported symptoms, albeit of different qualities and quantities (figure 4a). The most prominent symptom recorded by 69.23% of the NT-positive participants was anosmia/dysgeusia (figure 4b). However, the number of symptoms did not correlate with the level of the neutralisation titre (figure 4c). Of notice was that even in those with risk factors and neutralising antibodies, indicating past infection, neither a severe course of disease nor hospitalisation was reported. Regarding age distribution, the highest number of NT-positive test results was found in those aged 25 to 50 years, with slightly more males affected (53.85%) (figure 4d and e).

**Figure 4:**
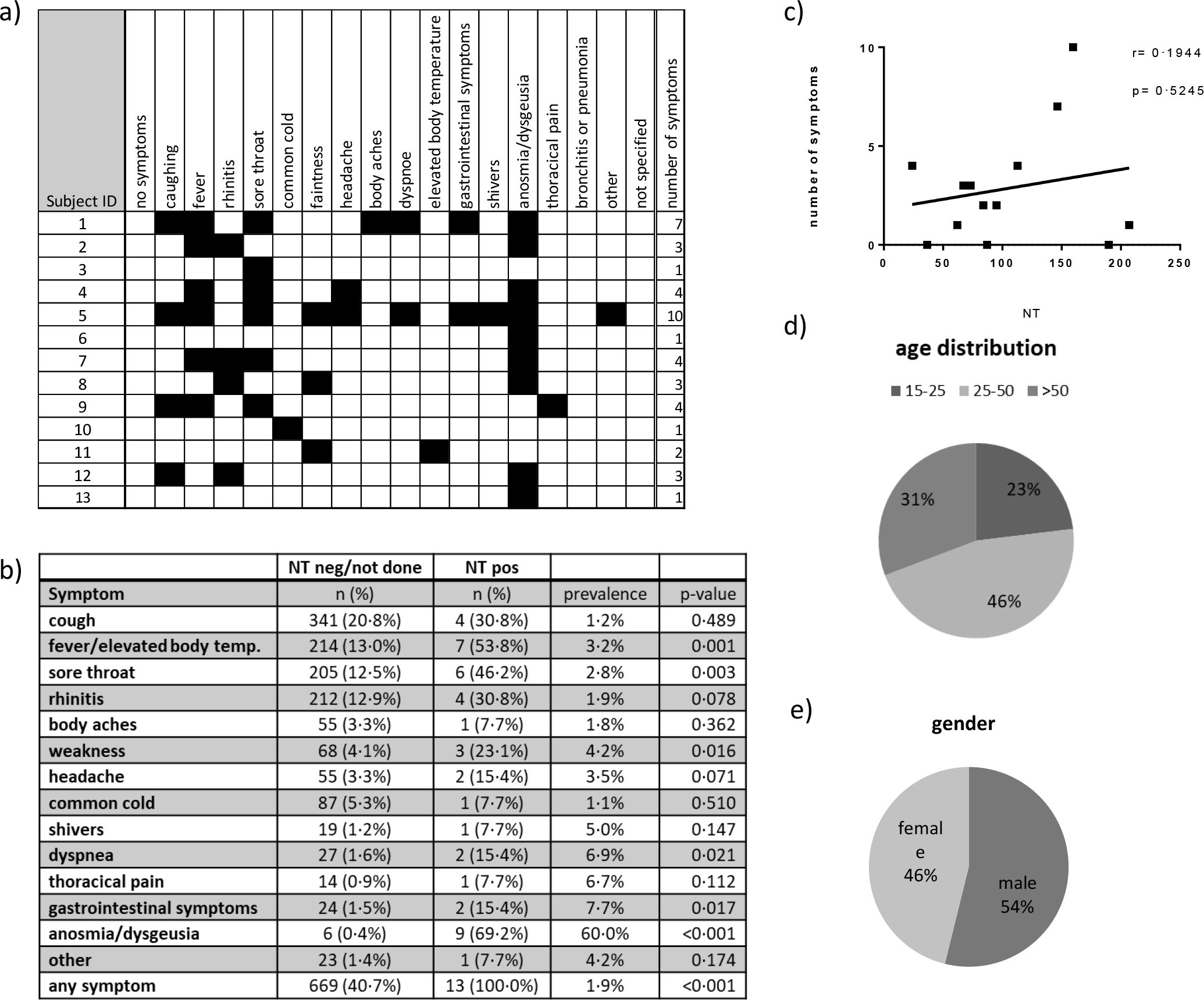
Individual symptoms (a), overall prevalence of COVID-19 like sympotms in participants without and with neutralising antibodies (b), correlation of the the number of symptoms with the neutralisation test (NT 1:X) result (c), distribution with regard to age group (d) and gender (e) among participants with neutralising antibodies evaluated in sera from the initial blood draw (day 0).

### Antibody persistence

We were highly interested in exploring the longevity of robust seropositivity, i.e. detection of protective/neutralising antibodies, in our study population. Therefore, all participants with detectable virus-reactive antibody levels (n=168) were invited for further blood draws after three and six months. Again, antibodies binding to S1, RBD, and NCP were measured. Only the RBD-specific total antibody levels, which highly correlated with the neutralising antibodies, showed stable persistence, indicating that protective antibodies maintained for at least six months (figure 5). RBD-specific IgM was lost in 53.33% at already three months and remained relatively stable thereafter. In contrast, S1-specific IgG and IgA antibody levels, which may indicate virus contact but not necessarily protection, rather tended to decline within three months in 37.5% and 53.68% of the participants, respectively (figure 5). Nevertheless, when S1-specific IgG antibodies were detectable at three months, they further persisted in 90% of the cases (9/10), also up to six months. NCP-specific IgG was lost in 9.09% at three months, whereas 55.55% of NCP-specific IgG were lost at six months. Measurements of the total NCP-specific antibodies confirmed the result of the NCP-specific IgG assay and also showed that the antibodies remained positive for three months (figure 5).

**Figure 5.**
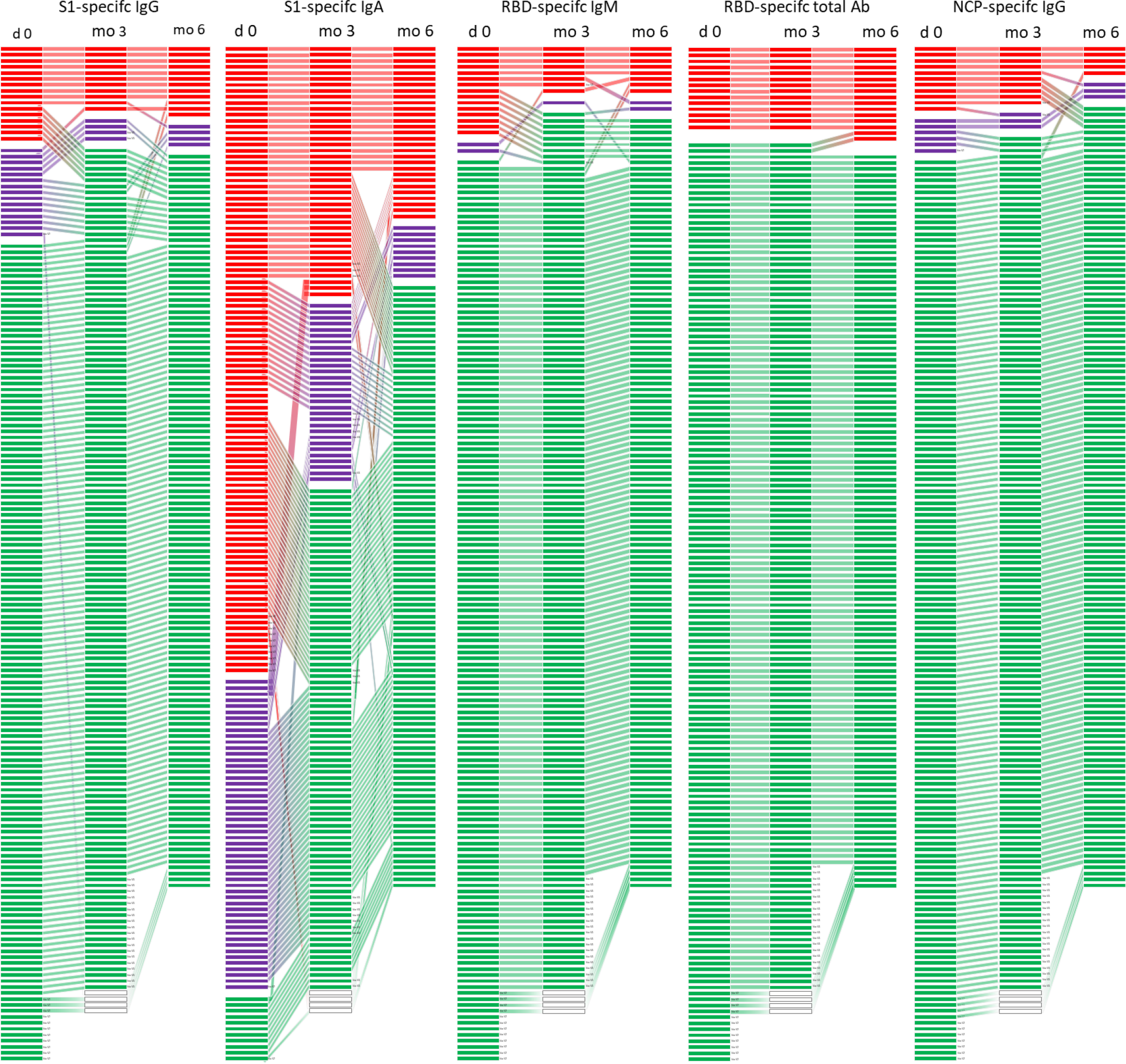
Development of individual antibody results at three different time points (day 0 (n=168), at three (n=152) and six months (n=139)) measured against different SARS-CoV-2 antigen specificities (S1, RBD and NCP) in the subgroup of those with detectable S1-reactive antibodies at the first blood draw. Each line represents one participant, red lines represents a positive result, violet a borderline result and negative results in green.

### Changes in seroprevalence over six months

At six months, all participants were invited again to assess how seroprevalence evolved. In total, we could evaluate antibody levels from 78.07% (1292/1655) volunteers, showing a non-significantly increased ratio of newly positives to those that became negative (chi^2^ McNemar p=0.212) for S1-reactive IgG (table 3). The prevalence of S1-reactive IgA antibodies decreased (p > 0.001) compared to the initial blood draw. RBD IgM antibody positivity as well as NCP IgG positivity increased over the six month period, although about 50% of previously positives became negative. With regard to the RBD-specific total antibodies none of the previously positive antibodies declined to negative concentrations, while 2.1% of the previously negative became positive. This increase in RBD seropositivity as well as the stability of these antibodies over a period of six months is expressed as ratio > 54 (table 3).

**Table 3:** Changes in the seroprevalence between day 0 and six months indicated by the ratio of newly positives to those that became negative; and the proportion of negatives that became positive (neg->pos) as well as the proportion of positives that became negative (pos->neg) and difference of prevalences of positives at month 6 and day 0; p-value assessed by Chi^2^ McNemar·

| Test | ratio | p-value<br>(against day 0) | neg->pos<br>(%neg) | pos->neg<br>(%pos) | Difference of<br>prevalence (%) |
| --- | --- | --- | --- | --- | --- |
| S1-specific IgG | 1.56 | 0.212 | 2.0 | 55.2 | 0.7 |
| S1-specific IgA | 0.33 | <0.001 | 2.6 | 70.8 | -4.8 |
| RBD-specific IgM | 2.88 | 0.012 | 1.8 | 53.3 | 1.2 |
| RBD-specific<br>total antibodies | >54 | <0.001 | 2.1 | 0.0 | 2.1 |
| NCP-specific IgG | 3.00 | 0.014 | 1.6 | 50.0 | 1.1 |

## Discussion

In our longitudinal study, we aimed to evaluate the seroprevalence, seroprotection, and duration of immunity against SARS-CoV-2 in a representative cohort of 1655 working adults over at least six months. An important aspect was to investigate which serological assay with respect to antigen specificity would be most appropriate for large-scale screening of past infections and seroprotection. Furthermore, we intended to investigate whether specific symptoms may serve as prediction markers of seropositivity and whether specific demographic parameters or working circumstances influenced the likelihood of virus contact/infection. Finally, we also aimed to explore the duration of antibody responses and seroprotection up to six months.

Our study population comprised of three age groups ranging from 16 to 65 years representative of the Austrian adult working population. For age, logistic regression analysis showed that the likelihood of seropositivity decreased with age. At the initial blood draw, antibody screening was performed using one of the first tests on the market, a S1-specific ELISA. Of all participants, 10.15% had IgG and/or IgA antibodies (above the cut-off, including borderline values) with 9.52% of these being positive for S1-specific IgG. These results correspond to those of a preceding Austrian study, in which 1544 random PCR samples were tested between the 1^st^ and the 6^th^ of April 2020, showing that the maximum prevalence of infected individuals was 0.33% (upper 95% confidence interval value).^7^ Our results of S1-specific IgG antibodies being prevalent in 0.97% of all participants would refer to the accumulated number of cases until that time. Similar rates have been reported for population-based seroprevalence studies in other European countries, while the rates have increased over time and varied substantially (0.4% to 14%) within different geographical regions relating to the occurrence of major infection clusters.^8,9^

At the beginning of the pandemic and throughout the Austrian pre-lockdown period, the employer facilitated home office work for the majority of their employees, particularly those of higher age and with potential risk factors for severe COVID-19. A certain number of employees, mainly the younger individuals, continued to work on-site during the overall study period, presumably exposing them to a higher risk of viral infection than those working from home. It could be due to this fact that the majority of the youngest group of 15-to 25-year-old employees, therefore, had the highest contact rates with colleagues or costumers, as this age group showed the highest levels of seropositivity. The percentage of S1-specific antibodies was lowest in the oldest age group, possibly explained by adequate compliance with the recommended hygiene measures and contact restrictions. Surprisingly, a higher prevalence of S1-specific antibodies was found in the group of participants working from home despite correction for age. However, this observation may in part be attributed to a bias related to the company’s policy that all employees with respiratory symptoms should stay at home.

Evaluation of the recent medical histories revealed that 42% of all participants had experienced respiratory symptoms, such as cough, fever, sore throat, and rhinitis. Concurrent circulation of other respiratory infections during winter and early spring could explain the fact that only some of them had detectable antibodies against S1 (9.91%). Notably, while the overall appearance of symptoms did not correlate with seropositivity, anosmia, or dysgeusia can be regarded as predictive markers for infection with SARS-CoV-2 and subsequent seropositivity.

On the other hand, the relatively low number of S1 antibodies in relation to the recorded symptoms may be ascribed to a fast waning of this antibody type, as was recently described by other authors.^10^ Furthermore, it was previously reported that approx. 10% of (non-hospitalised) patients presenting with mild COVID-19 did not mount detectable S1 antibody responses.^11,12^ Thus, as we do not have PCR results from all our participants with recorded symptoms, some mild infections may have been missed in our study as well.

The improvement of the antibody test systems enabled detection of antibodies directed against different antigenic regions, such as the RBD and the NCP. Recent studies have indicated that the severity of symptoms/disease has an impact on antibody levels and possibly also on their specificities.^5,11-13^ Along these lines, we tested the antibody responses for these additional SARS-CoV-2 antigens within the group of participants, who had been identified as S1-seropositive. Only 15.5% of the sera from participants with S1-specific antibodies also displayed antibodies directed against RBD or NCP. Notably, S1-specific IgA ratios, even at high levels, did not correspond with RBD- and NCP-specific antibody levels. This may be because the specificity of this test varies between 73%^14,15^ and 94%, resulting in a very low estimated positive predictive value of approx. 39% according to Gereuts van Kessel et al.^16^ The low specificity of SARS-CoV-2-specific IgA detection may be due to unspecific binding or cross-reactions of antibodies with other respiratory viruses.^14^ Additionally, the role of serum IgA in COVID-19 disease is not yet clear. The very high IgA titres may still follow contact with the virus and could represent the mucosal activity of dimeric IgA – however, this needs to be further confirmed.^12^ Thus, the overall lower specificity of both IgA and IgG antibodies against S1, as reviewed recently,^17^ also suggests that S1-specific antibody ELISA is not the optimal test for seroepidemiological surveys in low-prevalence settings (<5%) due to the reduced positive predictive value.^4,18^

Importantly, we wanted to test whether the antibodies measured were also associated with protection. In this respect, neutralising antibodies are regarded as surrogate marker of protection.^19^ According to an experimental model with macaques, neutralising antibodies against SARS-CoV-2 may play an essential role in protection against reinfection.^20^ In our study, the total RBD antibodies showed the highest correlation (kappa = 0.9448) with the neutralising antibodies of a live virus assay. Our data are supported by a recent study indicating that virus neutralisation is linked to B cell epitopes of the S protein, in particular neutralising epitopes of its RBD.^21,22^ In contrast to approx. 19% of S1-positive but asymptomatic participants, all of the participants with neutralising antibodies also showed symptoms – even if some had only one or two mild symptoms of which anosmia or dysgeusia was the most prevalent. Thus, anosmia or dysgeusia may even be regarded as a highly reliable diagnostic marker in very mild cases, as also proposed by other colleagues.^23,24^ Of note, we could not find a correlation between the number of symptoms (implying severity) and the level of the neutralisation titre as supported by other recently published data.^25,26^ Our data further indicate that in individuals with anosmia/dysgeusia as a sole symptom, the quality and quantity of neutralising/protective antibodies does not differ compared to those with several reported and typical COVID-19 symptoms (fever, cough, dyspnoea). Thus, our observation does not confirm suggestions that people with mild symptoms do not develop robust protective/neutralising antibody responses.^5,27^

Of major importance is the duration of protective immune responses. A recent study in COVID-19 patients and patient contacts showed that S1-specific and neutralising antibodies could last for up to five months.^28^ In contrast, a study in healthcare workers, though in a rather small cohort, postulated that the virus-specific antibody responses to the S antigen are only of short duration, in particular in individuals with mild or asymptomatic courses of disease.^29^

By analogy with the study authored by Patel et al., the results of our investigation indicate that a high percentage of the S1-specific IgG and IgA antibodies declined already after three months. Similarly, the NCP-specific antibodies also started to decline after three months. The drop in S1-specific antibodies was most evident and may be explained by an asymptomatic or oligosymptomatic course of the disease. Of interest, we here show that the kinetics of antibody decline differed according to antibody specificity over time. While the S1-specific antibodies decreased mostly within the first months, the NCP-specific IgG was solidly detectable for three months and thereafter declined until up to six months. In total contrast, the RBD-specific antibodies, correlating with the protective/neutralising antibodies, were consistently stable up to six months, and the number/severity of symptoms did not affect the duration of seroprotection. These results in mind, testing for RBD-specific antibodies should deliver the most reliable results to determine seroprevalence up to several months after infection. In contrast, cases may be lost already after three months when evaluating S1- or NCP-specific antibody response. Considering data from the previous SARS-CoV-1 pandemic, during which neutralising antibodies remained detectable in most patients for two years, it can be assumed that the protective/neutralising antibodies will persist over the next months.^30^

With the observation time of six months, we can also analyse changes in seroprevalence. Our results of the total study population show that in addition to the 0.85% RBD-positive participants at the initial blood draw, 2% additionally became positive after 6 months, thus reflecting the recorded increase in cases in Austria during the autumn of 2020. Further evaluation covering up to one year is planned.

In summary, we report a low seroprevalence of 0.97% with regard to S1-specific IgG antibody levels within the adult population at the early beginning of the pandemic. However, for immunity and protection, i.e. detection of neutralising/protective antibodies, the percentage is even lower with 0.85%. Importantly, these protective antibodies last for at least six months irrespective of a mild/oligosymptomatic or polysymptomatic course of the disease. Of all clinical symptoms, anosmia/dysgeusia was the most reliable symptom which was also associated with the generation of robust neutralising antibodies. In contrast, respiratory symptoms were not reliable diagnostic markers to predict antibodies or protection in the total study population. Large-scale seroprevalence studies can benefit from the use of a screening test with high SARS-CoV-2 specificity, as shown, RBD-specific assays could reliably detect SARS-CoV-2 specific antibodies for at least six months. Eventually, positive results may be retested with an appropriate neutralisation test to confirm the protective capacity of these antibodies.

## Data Availability

All data are available upon request.

## Acknowledgements

We would like to thank Sylvia Rudolf, Romana Hricova, Karin Baier, Maria Orola, Doaa al Mamoori, Tatjana Matschi, Vanessa Maurer, Barbara Schaar, Karin Schoiswohl, and Andrea Wendl for their excellent administrative and technical support. In addition, we would like to thank Thomas Perkmann (Laboratory Medicine, Medical University of Vienna) for the total NCP antibody measurements with the Roche Elecsys® assay. The contributions of Melanie Graf, Brigitte Kainz and Julius Segui (neutralisation assays) are gratefully acknowledged. SARS-CoV-2 was sourced via EVAg (supported by the European Community) and kindly provided by Christian Drosten and Victor Corman (Charité Universitätsmedizin, Institute of Virology, Berlin, Germany).

## Author contributions

Study conception and design: U.W., E.H., M.K.

Development and methodology: A.W., A.G., J.J., U.W, H.S., M.R.F., T.R.K., M.K.

Collection of the data: A.W., A.G., J.R., J.J., U.S., I.Z., M.R.F., T.R.K.

Data analysis and interpretation: A.W., A.G., J.J., U.S., M.K., H.S., M.F., T.K., U.W.

Writing all sections of the manuscript: A.W., A.G., U.W.

Manuscript revision: A.W., A.G., J.R., J.J., U.S., I.Z., M.K., H.S., M.R.F., T.R.K., E.H., U.W.

## References

1. Hartley DM, Perencevich EN. Public Health Interventions for COVID-19: Emerging Evidence and Implications for an Evolving Public Health Crisis. JAMA 2020.

2. WHO. Considerations for public health and social measures in the workplace in the context of COVID-19. 2020.

3. Koopmans M, Haagmans B. Assessing the extent of SARS-CoV-2 circulation through serological studies. Nat Med 2020; 26(8): 1171–2.

4. Bryant JE, Azman AS, Ferrari MJ, et al. Serology for SARS-CoV-2: Apprehensions, opportunities, and the path forward. Sci Immunol 2020; 5(47).

5. Poland GA, Ovsyannikova IG, Kennedy RB. SARS-CoV-2 immunity: review and applications to phase 3 vaccine candidates. Lancet 2020; 396(10262): 1595–606.

6. Vabret N, Britton GJ, Gruber C, et al. Immunology of COVID-19: Current State of the Science. Immunity 2020; 52(6): 910–41.

7. Ogris G. Spread of SARS-CoV-2 in Austria - PCR tests in a representative sample SORA Institute for Social Research and Consulting, 2020.

8. Pollan M, Perez-Gomez B, Pastor-Barriuso R, et al. Prevalence of SARS-CoV-2 in Spain (ENE-COVID): a nationwide, population-based seroepidemiological study. Lancet 2020; 396(10250): 535–44.

9. Stringhini S, Wisniak A, Piumatti G, et al. Seroprevalence of anti-SARS-CoV-2 IgG antibodies in Geneva, Switzerland (SEROCoV-POP): a population-based study. Lancet 2020; 396(10247): 313–9.

10. Ibarrondo FJ, Fulcher JA, Goodman-Meza D, et al. Rapid Decay of Anti-SARS-CoV-2 Antibodies in Persons with Mild Covid-19. N Engl J Med 2020; 383(11): 1085–7.

11. den Hartog G, Schepp RM, Kuijer M, et al. SARS-CoV-2-Specific Antibody Detection for Seroepidemiology: A Multiplex Analysis Approach Accounting for Accurate Seroprevalence. J Infect Dis 2020; 222(9): 1452–61.

12. Yu HQ, Sun BQ, Fang ZF, et al. Distinct features of SARS-CoV-2-specific IgA response in COVID-19 patients. Eur Respir J 2020; 56(2).

13. Okba NMA, Muller MA, Li W, et al. Severe Acute Respiratory Syndrome Coronavirus 2-Specific Antibody Responses in Coronavirus Disease Patients. Emerg Infect Dis 2020; 26(7): 1478–88.

14. Jaaskelainen AJ, Kekalainen E, Kallio-Kokko H, et al. Evaluation of commercial and automated SARS-CoV-2 IgG and IgA ELISAs using coronavirus disease (COVID-19) patient samples. Euro Surveill 2020; 25(18).

15. Traugott M, Aberle SW, Aberle JH, et al. Performance of Severe Acute Respiratory Syndrome Coronavirus 2 Antibody Assays in Different Stages of Infection: Comparison of Commercial Enzyme-Linked Immunosorbent Assays and Rapid Tests. J Infect Dis 2020; 222(3): 362–6.

16. GeurtsvanKessel CH, Okba NMA, Igloi Z, et al. An evaluation of COVID-19 serological assays informs future diagnostics and exposure assessment. Nat Commun 2020; 11(1): 3436.

17. Espejo AP, Akgun Y, Al Mana AF, et al. Review of Current Advances in Serologic Testing for COVID-19. Am J Clin Pathol 2020; 154(3): 293–304.

18. Mathur G, Mathur S. Antibody Testing for COVID-19. Am J Clin Pathol 2020; 154(1): 1–3.

19. Muruato AE, Fontes-Garfias CR, Ren P, et al. A high-throughput neutralizing antibody assay for COVID-19 diagnosis and vaccine evaluation. Nat Commun 2020; 11(1): 4059.

20. Chandrashekar A, Liu J, Martinot AJ, et al. SARS-CoV-2 infection protects against rechallenge in rhesus macaques. Science 2020; 369(6505): 812–7.

21. Poh CM, Carissimo G, Wang B, et al. Two linear epitopes on the SARS-CoV-2 spike protein that elicit neutralising antibodies in COVID-19 patients. Nat Commun 2020; 11(1): 2806.

22. Seydoux E, Homad LJ, MacCamy AJ, et al. Analysis of a SARS-CoV-2-Infected Individual Reveals Development of Potent Neutralizing Antibodies with Limited Somatic Mutation. Immunity 2020; 53(1): 98–105 e5.

23. Dreyer NA, Reynolds M, DeFilippo Mack C, et al. Self-reported symptoms from exposure to Covid-19 provide support to clinical diagnosis, triage and prognosis: An exploratory analysis. Travel Med Infect Dis 2020: 101909.

24. Zahra SA, Iddawela S, Pillai K, Choudhury RY, Harky A. Can symptoms of anosmia and dysgeusia be diagnostic for COVID-19? Brain Behav 2020; 10(11): e01839.

25. Wang P, Liu L, Nair MS, et al. SARS-CoV-2 neutralizing antibody responses are more robust in patients with severe disease. Emerg Microbes Infect 2020; 9(1): 2091–3.

26. Wolfel R, Corman VM, Guggemos W, et al. Virological assessment of hospitalized patients with COVID-2019. Nature 2020; 581(7809): 465–9.

27. Long QX, Tang XJ, Shi QL, et al. Clinical and immunological assessment of asymptomatic SARS-CoV-2 infections. Nat Med 2020; 26(8): 1200–4.

28. Wajnberg A, Amanat F, Firpo A, et al. Robust neutralizing antibodies to SARS-CoV-2 infection persist for months. Science 2020.

29. Patel MM, Thornburg NJ, Stubblefield WB, et al. Change in Antibodies to SARS-CoV-2 Over 60 Days Among Health Care Personnel in Nashville, Tennessee. JAMA 2020.

30. Lin Q, Zhu L, Ni Z, Meng H, You L. Duration of serum neutralizing antibodies for SARS-CoV-2: Lessons from SARS-CoV infection. J Microbiol Immunol Infect 2020; 53(5): 821–2.

