## supplementary materials for "A longitudinal seroprevalence study in a large cohort of working adults reveals that neutralising SARS-CoV-2 RBD-specific antibodies persist for at least six months independent of the severity of symptoms"

Supplementary material:

### Methods:

#### Definition of symptoms and risk factors:

Symptoms such as coughing, dyspnoea, thoracic pain, sore throat, rhinitis, elevated body temperature, fever, shivers, limb pain, weakness, headache, dysgeusia and/or anosmia, and gastrointestinal symptoms as described for COVID-19 were recorded, in addition to medical risk factors including those predisposing for a severe course of COVID-19. The risk factors were defined according to the Austrian Ministry of Social Affairs, Health, Care and Consumer Protection (chronic lung/respiratory disease, chronic cardiovascular disease, active cancer, immunosuppression and immunosuppressive drugs, chronic kidney disease, chronic liver disease with liver failure, diabetes, and arterial hypertension).

#### Testing for SARS-CoV-2 specific antibodies

First, all sera were tested for SARS-CoV-2-specific immunoglobin (Ig) A and IgG antibodies using a commercial ELISA kit (Euroimmune®, Euroimmun Medizinische Labordiagnostika, Lübeck, Germany) according to the manufacturer's instructions. The antigen used in this semi-quantitative assay is the S1 domain of the SARS-CoV-2 S protein. The sera were diluted 1:101 before incubation. Results with a ratio below 0.8 were interpreted as negative, ratios between 0.8 and 1.1 as borderline and above 1.1 as positive. Samples within the borderline range and with ratios close to the cut-off of 0.8 or 1.1 (value from 0.7 to 1.2) were repeated in two independent tests and the geometric mean was used for the final result. Therefore, samples that were positive for IgG, regardless of their IgA result, were regarded as "IgG-positive", those with borderline values for IgG after two repetitions, regardless of their IgA result, as "IgG-borderline" and those negative for IgG but IgA-positive or IgA-borderline as "IgA-positive or -borderline" samples. No valid interpretation is currently possible in the case of isolated positive IgA findings. Samples negative for IgG and IgA were considered as "IgG- and IgA-negative".

Second, all positive and borderline samples were further evaluated for antibodies against the RBD of the S protein and NCP. The RBD-specific antibodies were determined using a commercial available ELISA for IgM and total antibodies (ab) (Beijing Wantai Biological Pharmacy Enterprise, Beijing, China) as described in the manufacturer's instructions, leading to borderline results when the ratio was between 0.9 and 1.1. Samples within the borderline range and with ratios close to the cut-off of 0.9 or 1.1 (i.e. between 0.8 and 1.2) were repeated in two independent tests and the geometric mean was used for the final result. NCP-specific IgG antibodies were tested by ELISA (Euroimmune®, Euroimmun Medizinische Labordiagnostika, Lübeck, Germany) applying the same criteria for calculating the results as for the S1 ELISA following the manufacturer´s instructions. In addition, we measured total NCP-specific antibodies in an automated sandwich electrochemiluminescence assay (Elecsys®, Roche Diagnostics, Mannheim, Germany; on a Cobas e 801). Results with a cut-off index of $\geq 1$ were regarded as positive.

Finally, all IgG-positive sera and those with high IgA levels (Euroimmune ratio > 4) as well as a random sample (n=20) of the seronegative sera were also tested/confirmed by a virus-neutralising assay to evaluate the rate of functional (seroprotective) antibodies. The SARS-CoV-2 NTs were done in cooperation with Takeda (Vienna, Austria).

**SARS-CoV-2 neutralisation assay**

SARS-CoV-2 neutralization (NT) testing was done similar as previously described.[1] Briefly, Vero cells (ATCC CCL-81) sourced from the European Collection of Authenticated Cell Cultures (84113001) were cultured in TC-Vero medium supplemented with 5% fetal calf serum, L-glutamine (2mM), nonessential amino acids (1x), sodium pyruvate (1mM), gentamicin sulfate (100 mg/ml), and sodium bicarbonate (7.5%). SARS-CoV-2 strain BetaCoV/Germany/BavPat1/2020 was kindly provided by the Institute of Virology at Charité Universitätsmedizin, Berlin, Germany. For the SARS-CoV-2 neutralisation assays, samples were serially diluted 1:2 and incubated with 100 tissue culture infectious dose 50% ($TCID_{50}$) of SARS-CoV-2 per well. The samples were subsequently applied onto Vero cells seeded in tissue culture plates and incubated for five to seven days, after which the cells were evaluated for the presence of a cytopathic effect and the SARS-CoV-2 neutralisation titre ($NT_{50}$), i.e. the reciprocal sample dilution resulting in 50% virus neutralisation, was determined using the Spearman-Kärber formula and reported as 1:X.

**Statistical evaluation**

For sample size considerations, we expected to compile data sets from approx. 800 employees continuing to work on-site with client contacts and some 700 home office workers. Based on the numbers of notified SARS-CoV-2 PCR-positive individuals in Vienna and an estimated number of 10% unreported cases, it was assumed that approx. 1% of the population had already had contact with the virus prior to the blood draw. Presuming that these figures would also apply to the employees included in the study, the effect size expressed as an odds ratio of 3 at a two-sided level of significance of 5% resulted in a statistical power of 77% to compare employees on-site and in home office (Fisher´s exact probability test).

The data were evaluated for the two groups (working on-site and from home) and the subgroups stratified for age (15 to 25 years, 25 to 50 and above the age of 50). Seropositivity as a dependent variable was evaluated in a general linear model for binominal counts (see supplementary materials for more details). The primary predictor variable was defined according to the current workplace (home office or continuing to work with client contacts). Age, the number of household contacts, and the presence of underlying disease served as co-variables. To assess the relationship between various symptoms and seropositivity, a stepwise procedure was applied entering all mentioned covariates in one step and including symptoms in further steps with a significance level of 5% for inclusion and 10% for exclusion. The analyses were done using SPSS 26 (IBM Corp., New York, NY, USA) and the graphics were prepared with GraphPad Prism 7 (Graphpad Software, Inc., San Diego, CA, USA) or Excel (Microsoft Excel 2010, Microsoft Corporation, Redmond, WA, USA).

For changes in seroprevalence between the first blood draw and at six months we calculated the ratio of new positives compared to new negatives applying the $Chi^2$ McNemar test and the difference in prevalences.

**Results:**

**Demographic data with regard to travel and social contacts**

About two thirds (67.37%) of the volunteers were living in Vienna and one third thus needed to commute to work. Almost one third (27.98%) were sharing households with children younger than 15 years of age. Half of the participants (49.61%) had been travelling within the last three months before the first blood draw, either within Austria (winter/skiing holidays) or abroad (suppl. figure 3). Regarding the use of public transport, 26.95% answered that they continued to use public transport during the lockdown period. Two thirds (61.39%) maintained social contacts during the lockdown period and 2.18% stated that they had been in contact with SARS-CoV-2-infected individuals. In this regard, only two of these respondents were PCR-tested thereafter and both were PCR-negative. In total, 19 participants (1.15%) had previously been PCR-tested for SARS-CoV-2 before the first blood draw, of whom three showed a positive PCR result.

References:


1. Schwaiger J, Karbiener M, Aberham C, Farcet MR, Kreil TR. No SARS-CoV-2 Neutralization by Intravenous Immunoglobulins Produced From Plasma Collected Before the 2020 Pandemic. *J Infect Dis* 2020; **222**(12): 1960-4.

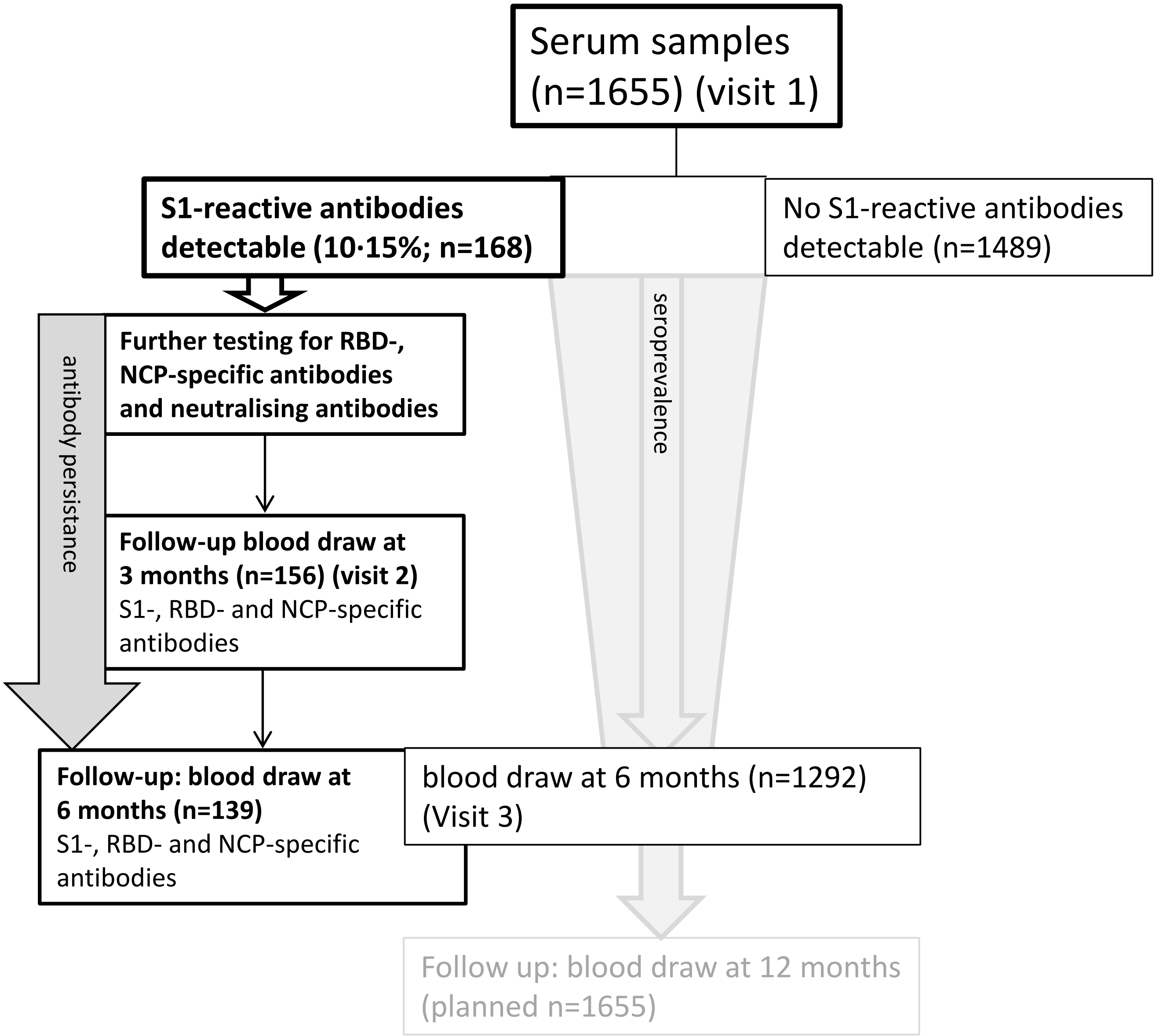


**Suppl. Figure 1: Flow chart of participants throughout the six months. Serum samples with S1-reactive antibodies at the initial blood draw and at six months were further analysed for RBD- and NCP-specific antibodies.**

a)

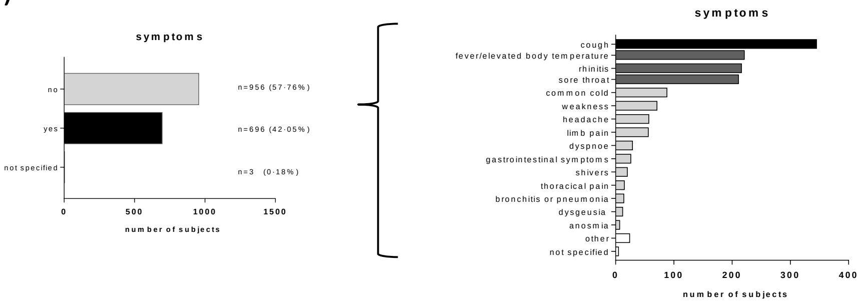


b)

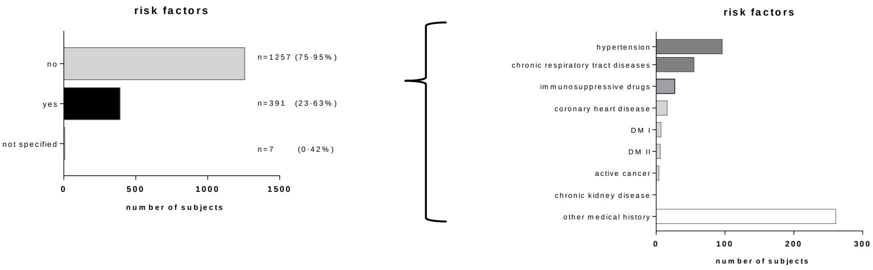


**Suppl Figure 2: Overall occurence of symptoms (a) within three months to the initial blood draw as well as presence of risk factors (b) the total study population that were reported at the day of the initial blood draw (day 0).** Multiple answers were possible

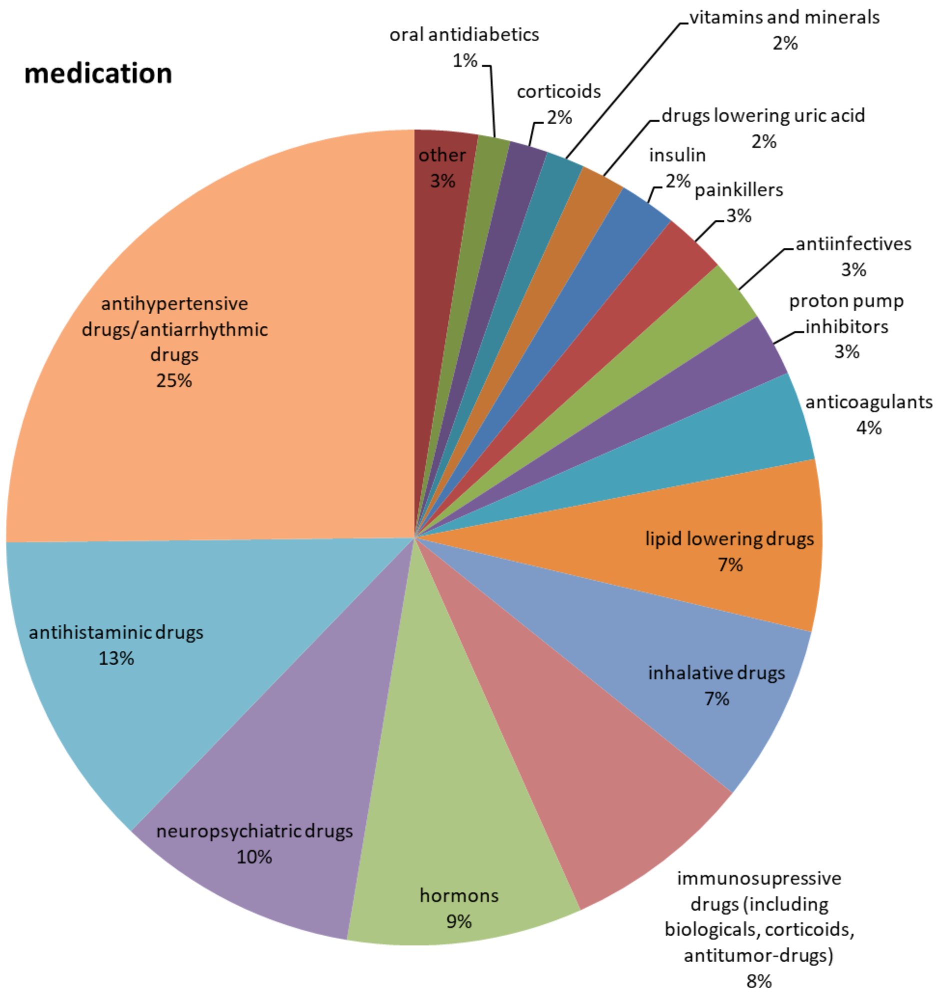


**Suppl Figure 3: Regular intake of medication as reported at the initial blood draw (day 0) from all participants.** Multiple answers were possible.

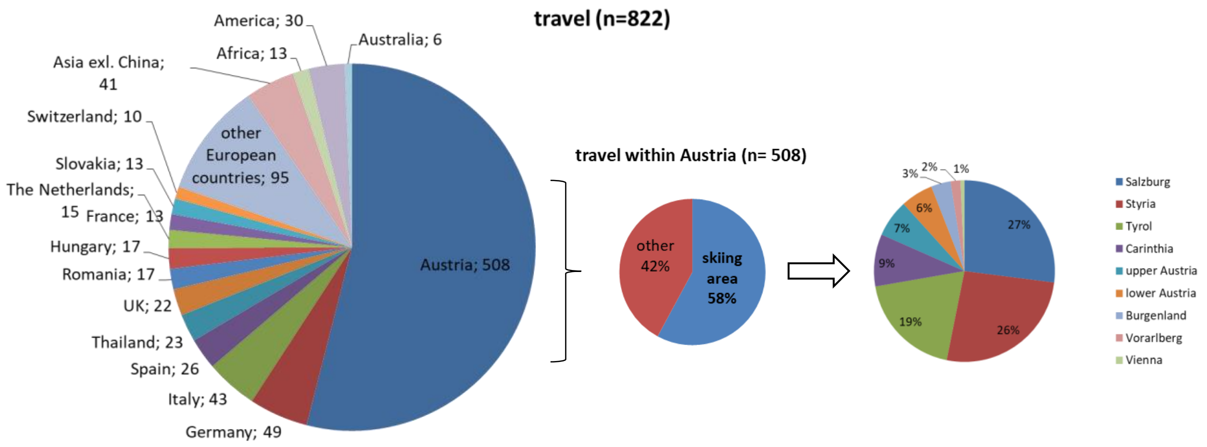


**Suppl Figure 4: Travel history within three months before the first blood draw as reported from all participants at the day of the initial blood draw (day 0).** Multiple answers were possible.

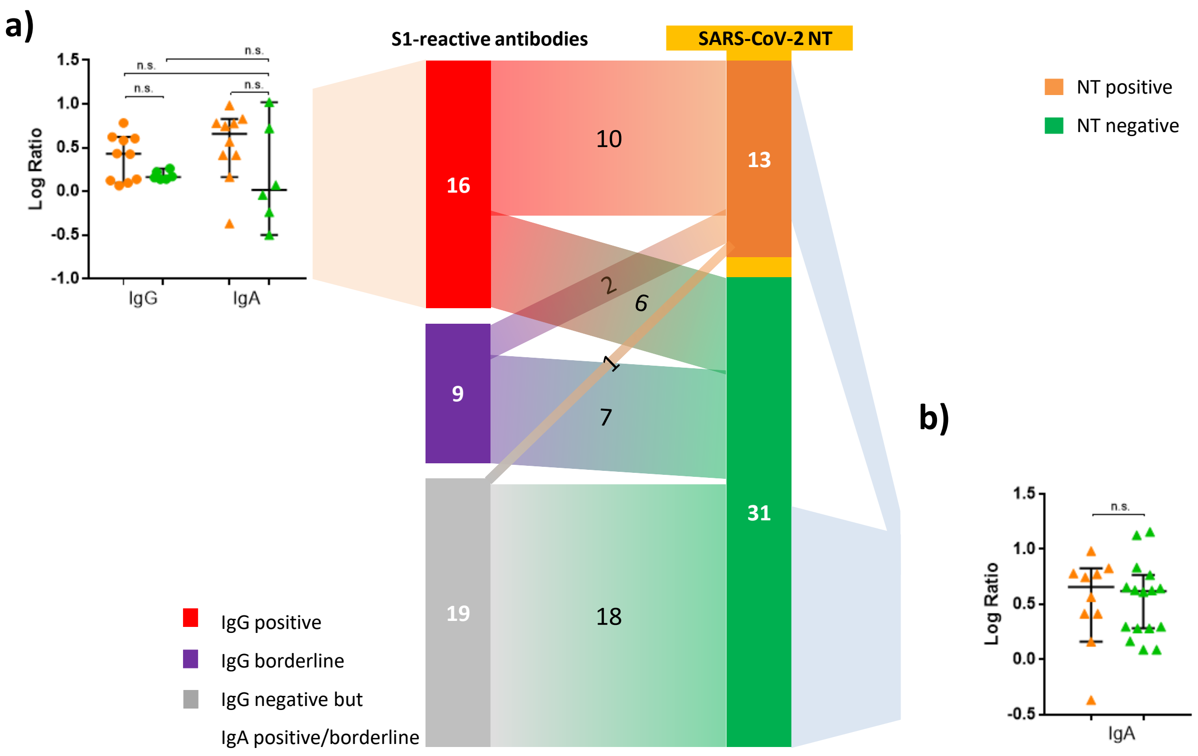


**Suppl Figure 5: Comparison of S1-reactive IgG and IgA antibody levels in participants according to the neutralisation test (NT) result (positive = orange; negative = green) in participants with a positive (red) S1-specific IgG result (a) and the comparison of the S1-reactive IgA ratio levels between those with a positive and a negative neutralisation test result at the first blood draw (day 0) (b).** S1-reactive antibody ratios presented after logarithmic transformation. Violet: borderline values for S1-reactive antibodies, grey: negative S1-reactive IgG but IgA positive/borderline.
